# Why has the United States of America not ratified the United Nations Convention on the Rights of the Child? The veto fulcrum as a new health policy analysis framework

**DOI:** 10.1101/2024.09.05.24312304

**Authors:** Lia Harris

**Affiliations:** Department of Pediatrics, University of British Columbia School of Medicine, Vancouver, British Columbia, Canada

**Keywords:** child health, health rights, global health policy, policy analysis framework

## Abstract

**Background:** The United Nations Convention on the Rights of the Child (CRC) enshrines health as a human right among other rights for children, fulfilled by each member state legally endorsing its principles through ratification of the Convention. Only the United States of America of all of the UN state parties has not ratified the CRC.

**Objective:** This study aimed to determine the reason(s) the CRC has not been ratified by the USA.

**Method:** Method design involved a mapping literature search and in-depth interviews with key stakeholders in the fields of global health, child rights, health policy, and US law. Implementing the health policy triangle systematized themes into content, context, processes, and actors as opposition or proponents of ratification.

**Results:** While most published opinions favour ratification of the CRC, critically, the literature focuses on pros and cons of ratification, rather than reasons why the US has not ratified. Interview informants expanded themes revealing a status quo of non-ratification which has become increasingly challenging to overcome. Drawing on veto player theory to explain the status quo and introducing a new policy analysis framework of a veto fulcrum reveals that within the process of ratification, single powerful actors at a veto fulcrum have made undemocratic decisions, obstructing CRC ratification.

**Conclusion:** This research has forged a new policy framework, the veto fulcrum, which examines political systems where political actors as veto players have extraordinary power to make executive decisions against public opinion, and against good health policy.

## Background

The United States of America (USA) Congressional webpage claims to put human rights at the center of foreign policy, yet it remains the exception on ratifying a human rights treaty which is meant to protect “the most voiceless, voteless, and vulnerable human beings on earth” [1,2]. It is essential, therefore, to investigate why America has not ratified the United Nations Convention on the Rights of the Child.

The 1989 United Nations Convention on the Rights of the Child (CRC or the Convention) was envisioned to assure the full rights and freedoms to which every person is entitled under the Universal Declaration of Human Rights, but with specific focus on children [3]. Human rights were enshrined at an international level in 1948 with the adoption of the Universal Declaration of Human Rights (UDHR) by the state parties to the United Nations at the time and have subsequently become part of legally binding international law, by means of two Covenants [4]. The UN member states later acknowledged the extra needs of children by drafting the CRC, beginning in 1979, the UN Year of the Child, adopting it in 1989 [3]. The CRC, meant to respect, protect, and fulfill the rights of children, has been the most rapidly and widely ratified human rights treaty in history.

Health is a central covenant in the CRC; borrowing the language from the Constitution of the World Health Organization (WHO), Article 24 of the CRC recognizes the rights of children to “the enjoyment of the highest attainable standard of health” [3,5]. Harnessing the universal recognition of child health as a human right within a global policy instrument should enshrine children’s entitlement to basic health care in not only wealthy countries, but critically for “children living in exceptionally difficult conditions, and that such children need special consideration” [3]. Therefore, even though the CRC is not specifically a health policy initiative, the universal consensus on the importance of child rights, including the acknowledgement of health as a human right for children, should be considered a first step in ensuring legal protection of health rights for children worldwide.

Ratification of the CRC signifies the endorsement of a member state on the importance of health and other rights for children within the state, and children of the world. Arguably, ratification does not guarantee respect for child rights by member states; indeed, violations of children’s rights continue to occur around the world, not the least of which is lack of health care [6]. However, ratification requires member states to submit reports to the Committee on the Rights of the Child two years after ratification then every five years thereafter, allowing the Committee to review and comment on states’ standing on child rights protection, and more importantly, giving civil society organizations opportunity to report violations [7,8]. Through these mechanisms, the CRC has been instrumentalized to help improve the health and wellbeing of children in ratifying member states. For example, ratification of the CRC was found to reduce child mortality in excess of natural trends following ratification by a member state [9].

### The USA and the CRC

Only one UN member state has not ratified - agreed to be legally bound by – the CRC. The USA became the outlier of 196 eligible UN member states in 2015 when Somalia and South Sudan were the last two states to accede to the Convention [3,10].

The United States was extensively involved in the drafting of the CRC. Drawn in part from the US Constitution, the United States made recommendations for 38 of the CRC’s articles and did ratify two optional protocols [10]. The US became a state signatory to the CRC on February 16, 1995, but has not subsequently ratified it, which requires the sitting American President to submit the treaty to the US Senate, who must approve it with a two-thirds vote [11]. Table 1 summarizes the process of international treaty ratification in the US [8].

**Table 1:**
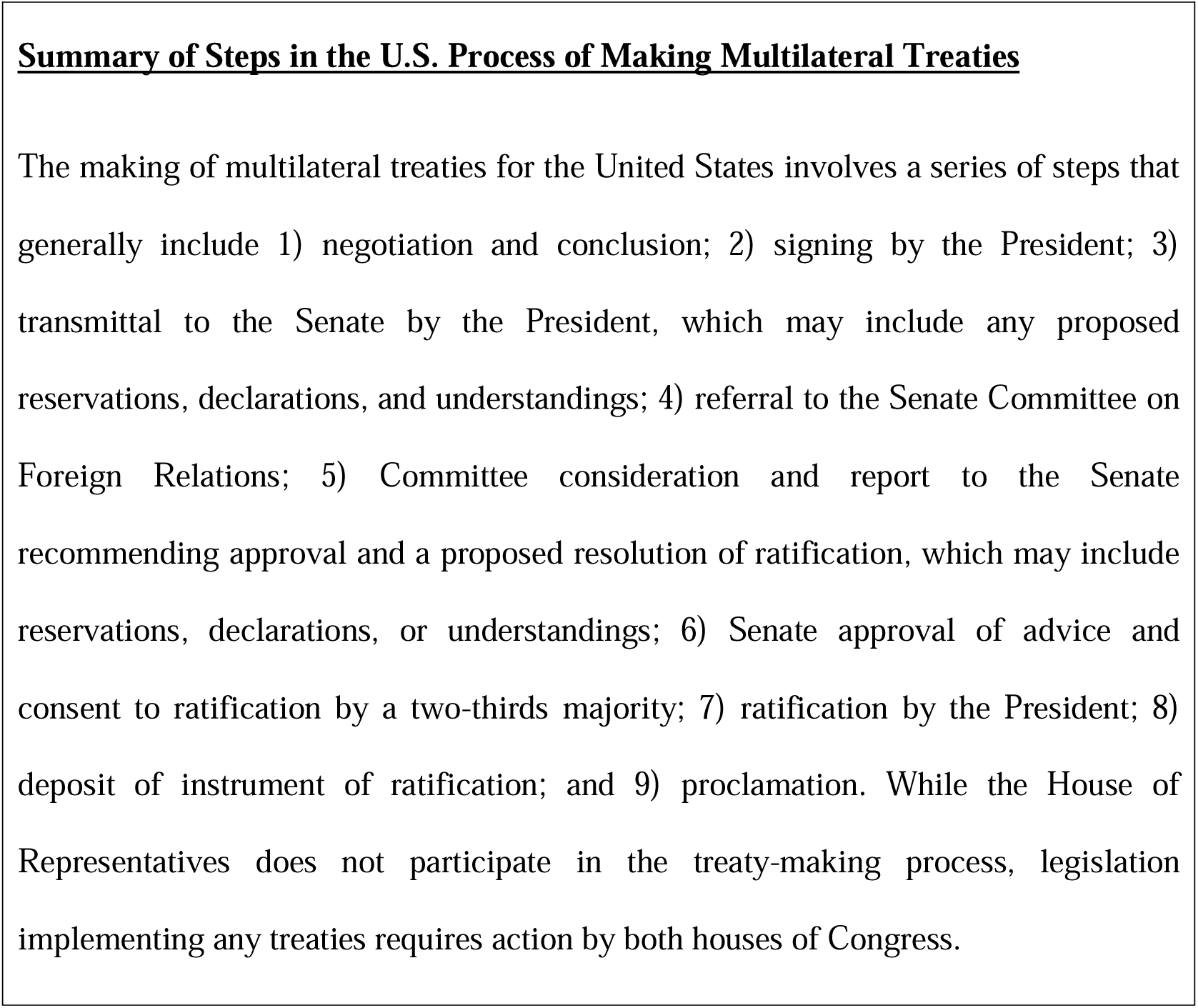
Summary of Steps in the US Process of making Multilateral Treaties from Blanchfield, L. Congressional Research Service. [8].

### Summary of Steps in the U.S. Process of Making Multilateral Treaties

The making of multilateral treaties for the United States involves a series of steps that generally include 1) negotiation and conclusion; 2) signing by the President; 3) transmittal to the Senate by the President, which may include any proposed reservations, declarations, and understandings; 4) referral to the Senate Committee on Foreign Relations; 5) Committee consideration and report to the Senate recommending approval and a proposed resolution of ratification, which may include reservations, declarations, or understandings; 6) Senate approval of advice and consent to ratification by a two-thirds majority; 7) ratification by the President; 8) deposit of instrument of ratification; and 9) proclamation. While the House of Representatives does not participate in the treaty-making process, legislation implementing any treaties requires action by both houses of Congress.

The CRC remains with the current President, to be put forth to the Senate for approval (step 3 in Table 1). The last president to address the issue was then-Senator, Barack Obama, who said it was “embarrassing” the USA had not ratified it, and had promised to “review this and other treaties to ensure the United States resumes its global leadership in human rights”, yet as President he did not submit it to the Senate [8,12]. Most recently, on February 12, 2020, Representative Ilhan Omar submitted Resolution 854 to the 116^th^ Congress, recommending the US ratify the CRC, which was forwarded to the House Committee on Foreign Affairs, but there has been no further activity [13].

Support for child rights in America and evidence for ratification of the CRC is overwhelming. A study conducted on the 20^th^ anniversary of the adoption of the CRC found that in a survey using random digit dialing of registered American voters, respondents favored ratification by a four to one margin (62-14%), and that opposition to ratification was low [14]. More recently, in two separate polls Americans indicate too little is spent on children and they support policies which would assist families with children, including policies involving tax credits, childcare, paid leave, adoption, health care, and housing [15,16]. Further, three quarters of Americans view health as a human right [17]. Ratification would encourage reforms which could reduce the inequity experienced by American children; the US falls behind other wealthy nations in multiple spheres of protection and provision for its children including poverty, health care, mortality rates, juvenile justice, education, labour laws, and child marriage [18–22].

### Policy Frameworks

Analysis and synthesis of the qualitative data was guided by a conceptual framework developed by Walt and Gilson from the field of health policy analysis [23]. They found analysis of health policies tended to place excess focus on the content of a policy, ignoring the context, processes, and actors related to policy action. As a result, they developed the health policy triangle as a model to understand the factors and their relationships in policy implementation [ibid]. This framework was chosen because - while it was devised specifically as a health policy analysis framework - the four categories effectively enable investigation of any policy under consideration. Figure 1 demonstrates the relationship of the content, context, process, and actors of policy enactment.

**Fig 1.**
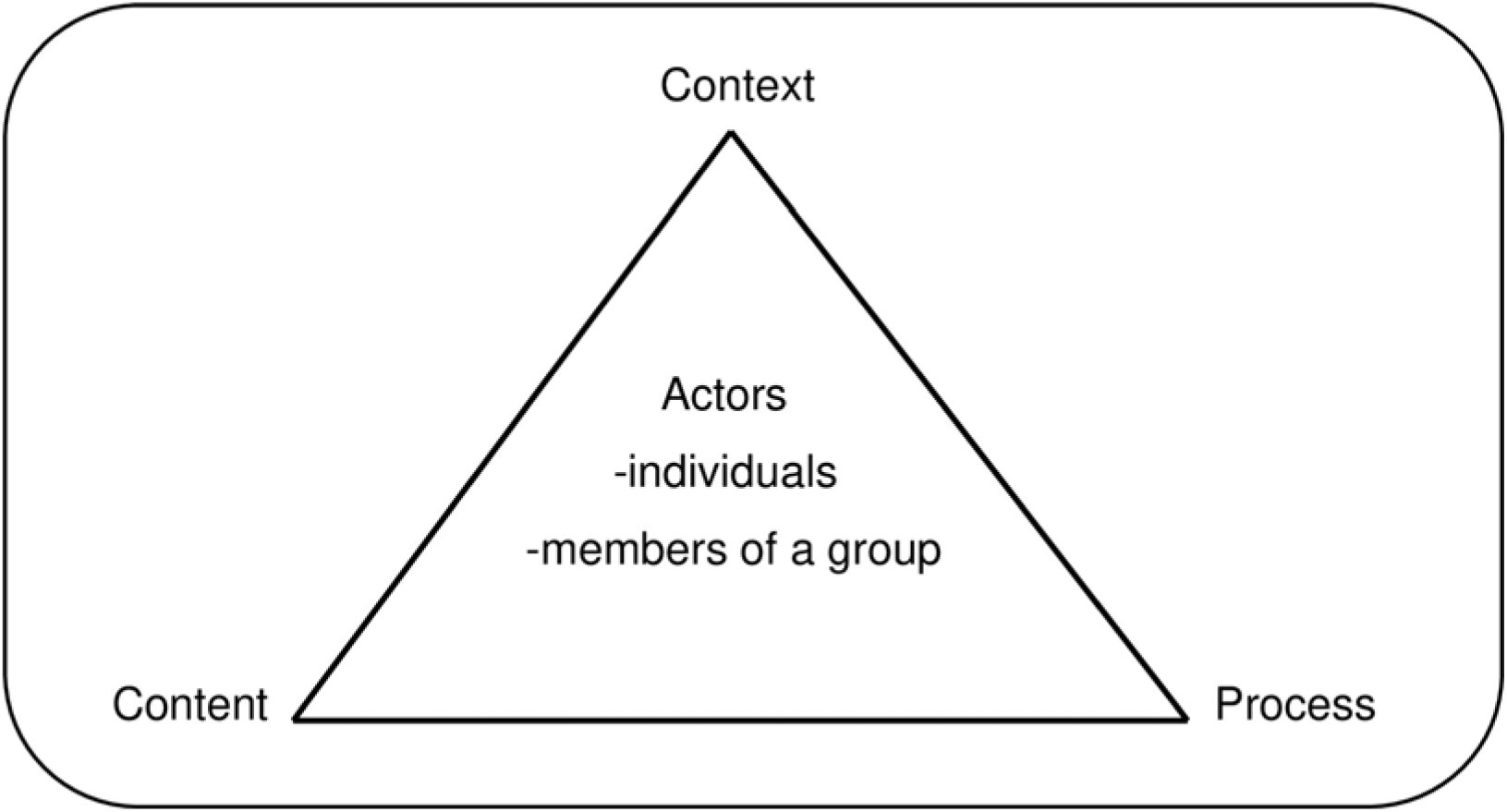
Health Policy Triangle from Walt and Gilson [23].

A complementary policy analysis framework, Tsebelis’ veto player theory, derives from political science, describing the inherent bias toward the status quo in policy making by identifying the key points in a political system at which policy initiatives can potentially be blocked [24]. A veto player is a political actor who can stop a change from the status quo [ibid]. Where there are a greater number of veto players, there is a greater chance of veto, and a tendency to the status quo. In this way, veto player theory highlights the dependent interaction between Walt and Gilson’s actor and processes regarding treaty ratification. This policy analysis framework was chosen to explore the role of specific actors - veto players - on the process of CRC ratification.

## Methods

### Study Design

The research question was approached using two methods, involving a mapping literature search and in-depth interviews with key stakeholders in CRC policy. A mapping review was chosen to enable systematic review of broader literature, because an initial scoping search failed to identify any primary research on the question. [25] Likewise, interviews were chosen as a supplemental secondary method because the literature did not answer why the US has not ratified the CRC. Interviews were conducted in accordance with the Declaration of Helsinki, the World Medical Association’s statement of ethical principles for research involving human participants.

### Stage 1: Literature Review

A systematic approach mapping review of the literature was conducted on four databases: Medline, Scopus, Web of Science, and Google Scholar for both published and grey literature since 1989 (chosen as the year of UN adoption) using the SPIDER search strategy tool [25–28]. The search terms and associated synonyms in combination with MeSH subject headings, where available, formed the search strategy for the literature review. Keyword searches included ‘United Nations Convention on the Rights of the Child’ or ‘CRC’ or ‘child rights’ and ‘United States of America’ or ‘USA’ and ‘ratify’ or ‘ratification’ or ‘agree’ or ‘agreement’ using Boolean operators. Non-systematic grey literature searches such as reference mining, citation searches, website searches, and searches of congressional records using the same search terms on the <u>congress.gov</u> website were included [26,28]. Grey literature such as press releases, news stories, editorials or blog posts were included, but evaluated as lower quality material. Articles were included in the study if they were published after 1989, English language, and related to USA ratification of the CRC. Publications which addressed US ratification of all of the UN human rights treaties, if they also included CRC, were included. Papers were included if they referenced CRC ratification by the USA, even if it was not the primary focus of the paper. The PRISMA flow diagram for databases, registers, and other sources was used to sort publications (Fig 2) [29].

**Fig 2.**
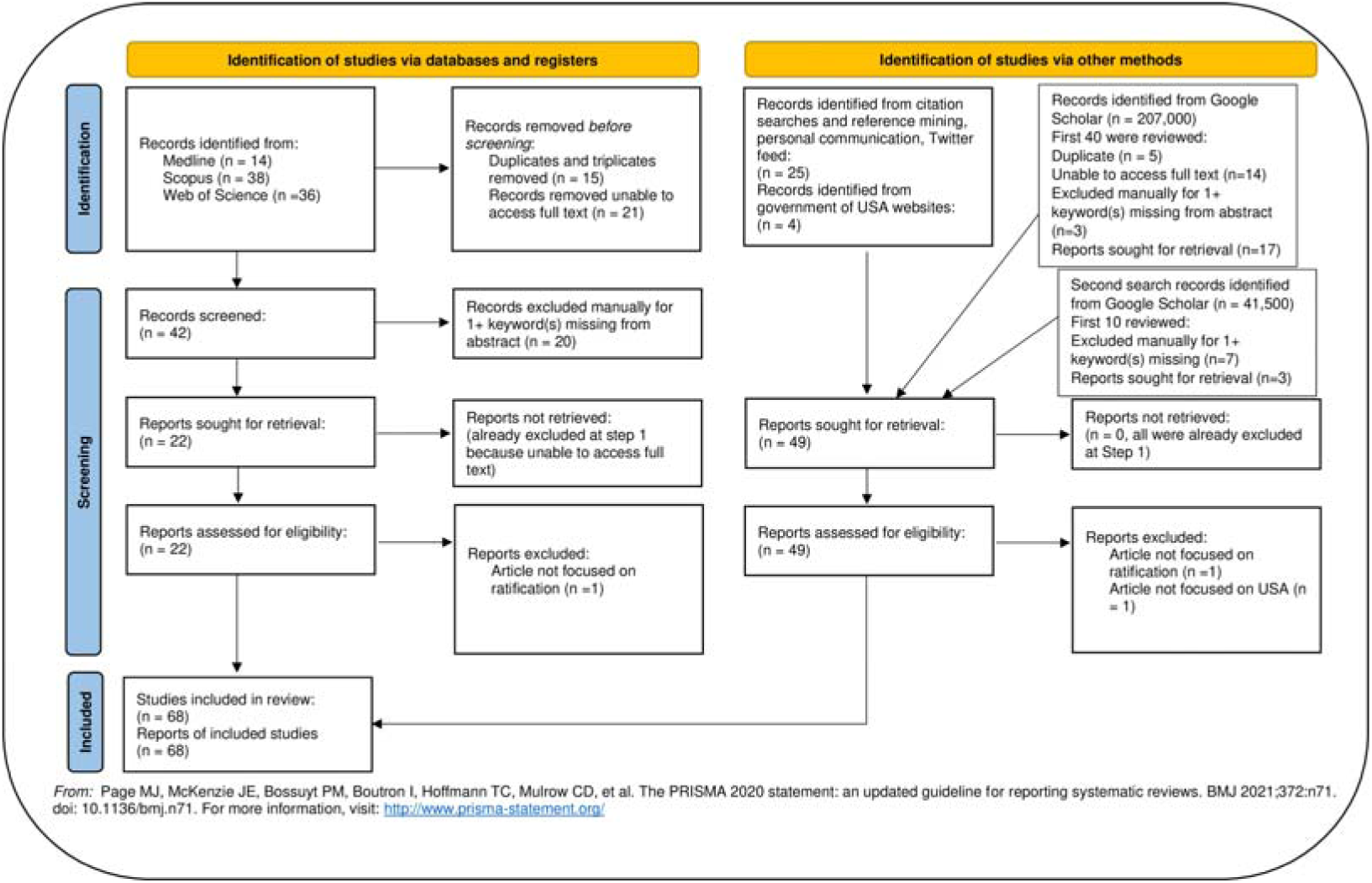
PRISMA flow diagram [29].

Articles obtained from the search were initially assessed for suitability from their titles and abstracts, then those found to be suitable were subjected to a review of the whole text.

### Stage 2: In-Depth Interviews

The second part of the research was conducting in-depth interviews with key informants who are stakeholders with respect to child health, child rights, and US policy law, who have professional knowledge of, or whose work relates to the CRC. Questions were semi-structured, loosely following the Interview Outline Questions informed by the literature search (Supplement 1). Some interview questions were standardized to ensure data source triangulation, to aim to ensure validity and reliability on data collected from different informants; however, the topic guide for the interviews was designed to be open-ended to maintain a sufficiently broad focus, to allow for hidden or emerging themes, and include questions specific to each informant’s area of expertise. On reflection of the bias inherent in interviewing only pro-ratification informants, invitations were extended to interview actors who oppose ratification. Selection of key informants was based on purposive sampling, by identification of individuals known to work in the global child health field or from literature searches, and then from snowball sampling/chain referral [30]. They were approached to participate through telephone, email, or social media, specifically Twitter and LinkedIn, depending on contact information available, using a standardized message. A starting number of 8-10 were chosen as possible informants for the in-depth interviews, but the final number of 14 informants was determined through achieving thematic saturation [31]. Inclusion criteria for possible informants were key stakeholders with knowledge of US law and/or treaty policy, child health policy, global child health, or child rights.

### Ethical Statements/Research Limitations

Informants were anonymized according to the Declaration of Helsinki, the World Medical Association’s statement of ethical principles for research involving human participants. The purposive and chain referral/snowball methods were limited by the primary researcher’s limitation and bias in access to informants, the most obvious omission being lack of access to key actors in CRC ratification – the US President and Senators. The single author status of the research should be considered a key limitation; while the author received input from a supervisor throughout the research, undoubtably, contributions and agreement among two or more authors would likely have enriched each step in the process, identified gaps, and produced more robust discussion. Limitations of the interviews might have included informants’ hesitation to give sensitive political opinions, and concern about whether the research aligns with their own priorities. Also, elite informants may have little time, and thus present standard responses in the interest of time [32].

### Data Analysis

1. In Stage 1, a thematic content analysis of the literature was conducted identifying themes relevant to the research question, particularly opposition themes related to content, context, processes and actors as described in Walt and Gilson’s health policy triangle. Data were extracted from the literature search and evaluated using a grounded theory approach both manually in Word and pdf documents with line-by-line open coding supported by NVivo to distil the broad patterns and consolidate them into themes [33,34]. Identified themes were classified into barriers/opposition and enablers/proponents of ratification, and sub-classified by content, context, process, and actors, according to the health policy triangle.

2. In Stage 2, thematic content analysis of the interview transcripts was conducted. Each interview was analyzed immediately following, with preliminary analysis informing further data collection in an iterative approach. Supporting the iterative approach, each informant was asked to suggest other potential informants. Themes derived from the interviews were compared with themes from the literature for external validity and representativeness. Interviews were evaluated using a modified version of the qualitative study CASP checklist (Supplement 2) [35]

## Results

### Stage 1: Literature Review

The literature review yielded 68 publications after PRISMA assessment for relevance (Fig 2). Most of the publications were not peer reviewed or systematic reviews. Rather, they were expert opinion or editorials in the fields of political science, US law, human rights, or public health policy. Four publications opined that the CRC should not be ratified (two from opposing actors, two indicating it would make no difference to child welfare), and eight others were neutral on the topic, three of which were congressional reports intended to be neutral and the other five suggesting ratification would not improve the wellbeing of children. Otherwise, 56 (82%) of the papers concluded with pro-ratification opinions. Nearly half (28/68, 41%) of the articles were from the discipline of US law, 12 each from medicine/public health and rights advocates, eight from political science, three from the field of psychology, and one each from the fields of anthropology, social work, religious studies, ethics, and education. Supplement 3 lists the publications and CASP evaluation.

### Generation of Themes from the Literature

Each publication was combed for themes which represented opposition and support to ratification using open coding [36]. The themes were thus organized into these eight subclasses, as shown in Table 2, with opposition themes defined in Tables 3 and 4.

**Table 2:**
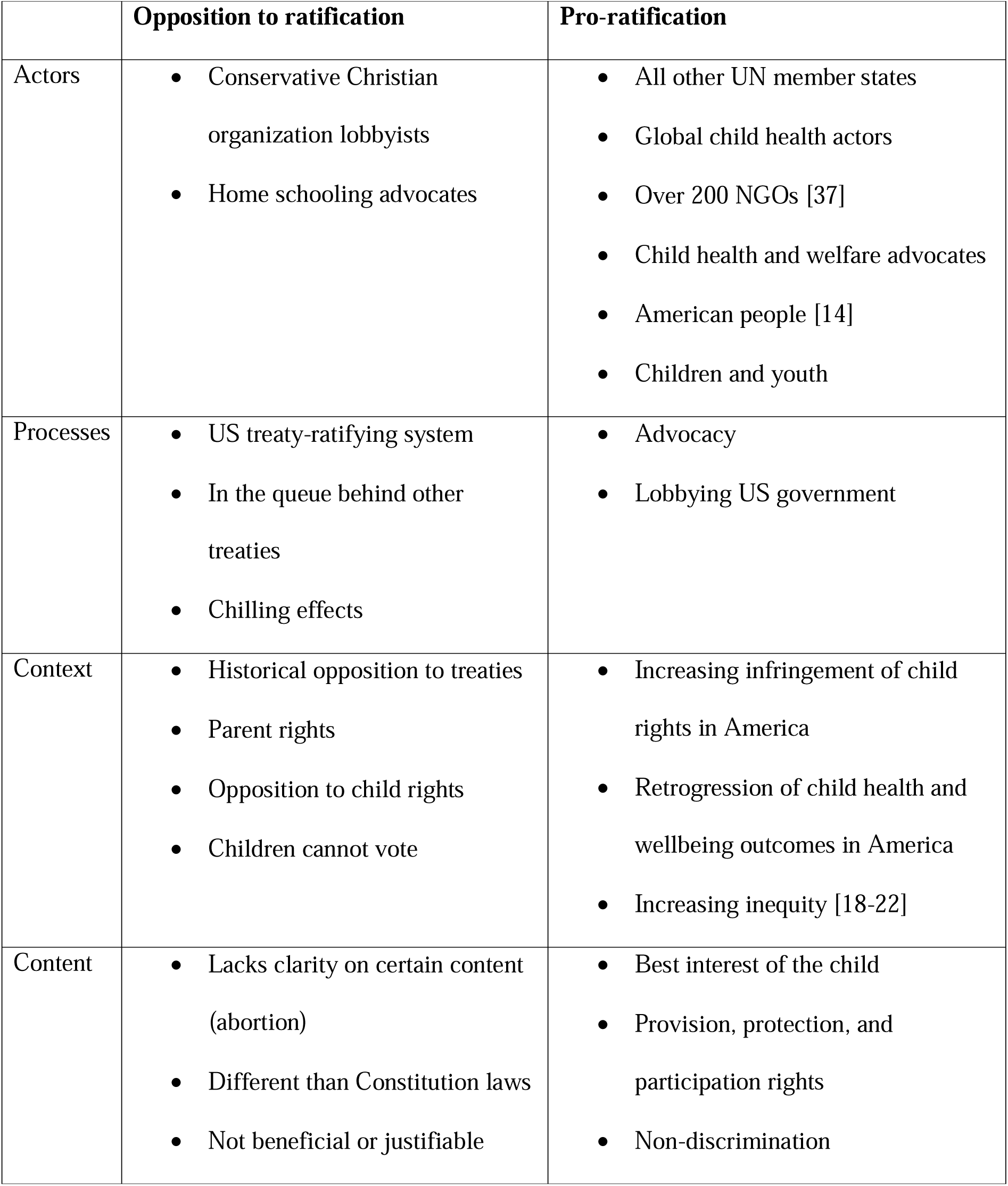
Opposition and Pro-Ratification Themes on US Ratification of the CRC in Walt and Gilson’s Health Policy Triangle.

|  | **Opposition to ratification** | **Pro-ratification** |
| --- | --- | --- |
| Actors | • Conservative Christian organization lobbyists<br>• Home schooling advocates | • All other UN member states<br>• Global child health actors<br>• Over 200 NGOs [37]<br>• Child health and welfare advocates<br>• American people [14]<br>• Children and youth |
| Processes | • US treaty-ratifying system<br>• In the queue behind other treaties<br>• Chilling effects | • Advocacy<br>• Lobbying US government |
| Context | • Historical opposition to treaties<br>• Parent rights<br>• Opposition to child rights<br>• Children cannot vote | • Increasing infringement of child rights in America<br>• Retrogression of child health and wellbeing outcomes in America<br>• Increasing inequity [18-22] |
| Content | • Lacks clarity on certain content (abortion)<br>• Different than Constitution laws<br>• Not beneficial or justifiable | • Best interest of the child<br>• Provision, protection, and participation rights<br>• Non-discrimination |

**Table 3:**
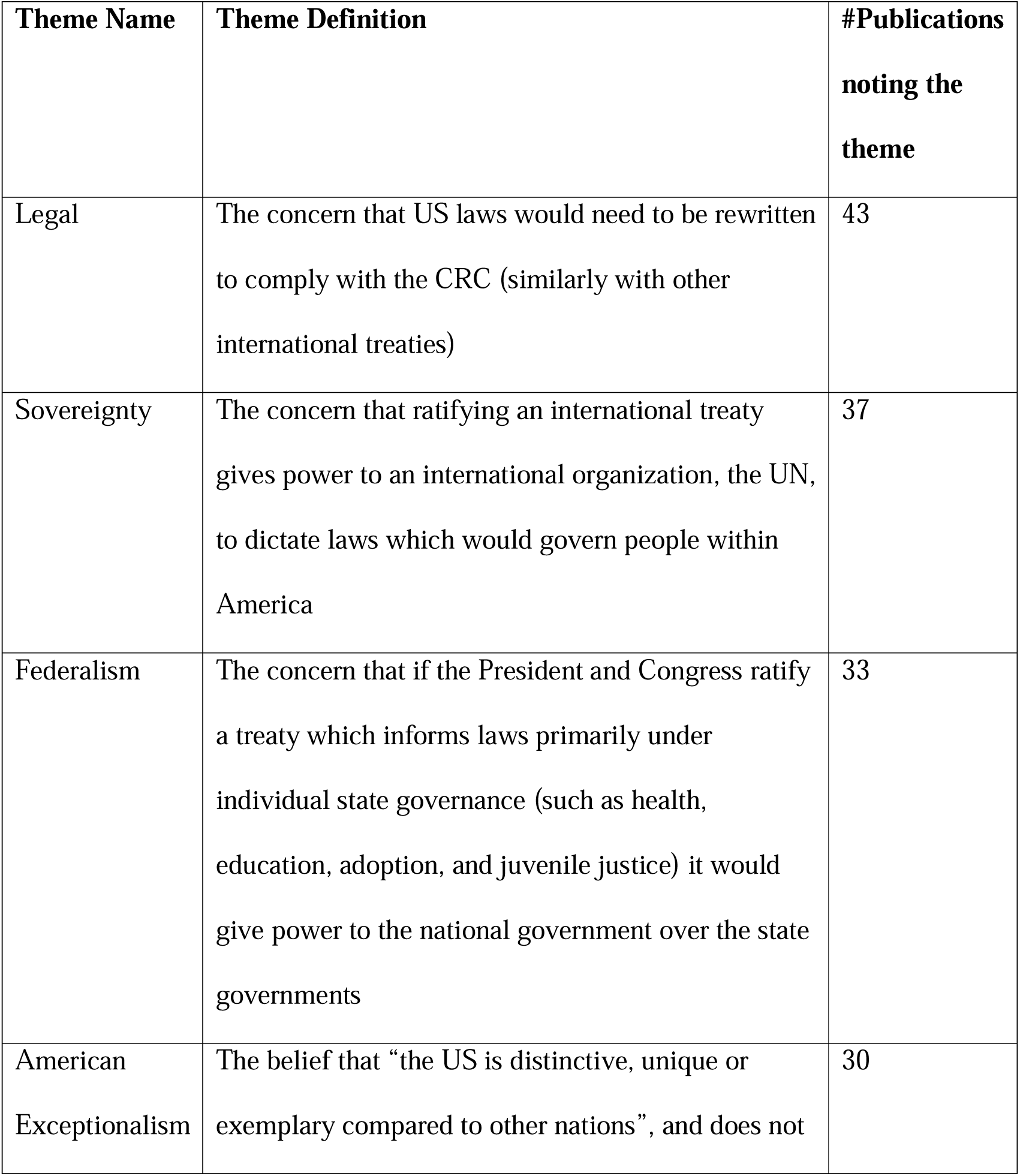
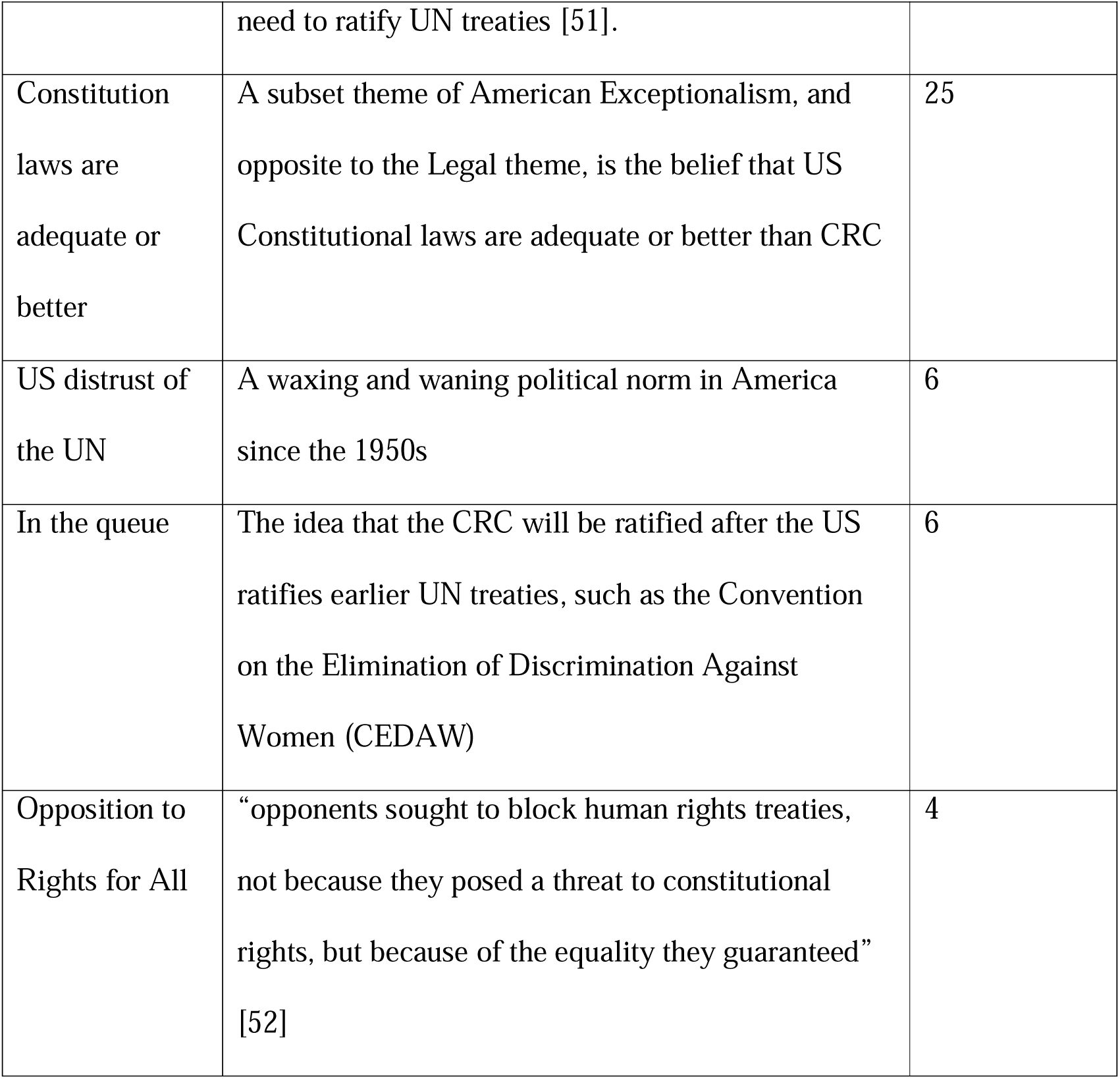
Historical Opposition to International Treaties (Context Themes)

| Theme Name | Theme Definition | #Publications<br>noting the<br>theme |
| --- | --- | --- |
| Legal | The concern that US laws would need to be rewritten to comply with the CRC (similarly with other international treaties) | 43 |
| Sovereignty | The concern that ratifying an international treaty gives power to an international organization, the UN, to dictate laws which would govern people within America | 37 |
| Federalism | The concern that if the President and Congress ratify a treaty which informs laws primarily under individual state governance (such as health, education, adoption, and juvenile justice) it would give power to the national government over the state governments | 33 |
| American<br>Exceptionalism | The belief that “the US is distinctive, unique or exemplary compared to other nations”, and does not | 30 |
|  | need to ratify UN treaties [51]. |  |
| Constitution<br>laws are<br>adequate or<br>better | A subset theme of American Exceptionalism, and<br>opposite to the Legal theme, is the belief that US<br>Constitutional laws are adequate or better than CRC | 25 |
| US distrust of<br>the UN | A waxing and waning political norm in America<br>since the 1950s | 6 |
| In the queue | The idea that the CRC will be ratified after the US<br>ratifies earlier UN treaties, such as the Convention<br>on the Elimination of Discrimination Against<br>Women (CEDAW) | 6 |
| Opposition to<br>Rights for All | “opponents sought to block human rights treaties,<br>not because they posed a threat to constitutional<br>rights, but because of the equality they guaranteed”<br>[52] | 4 |

**Table 4:**
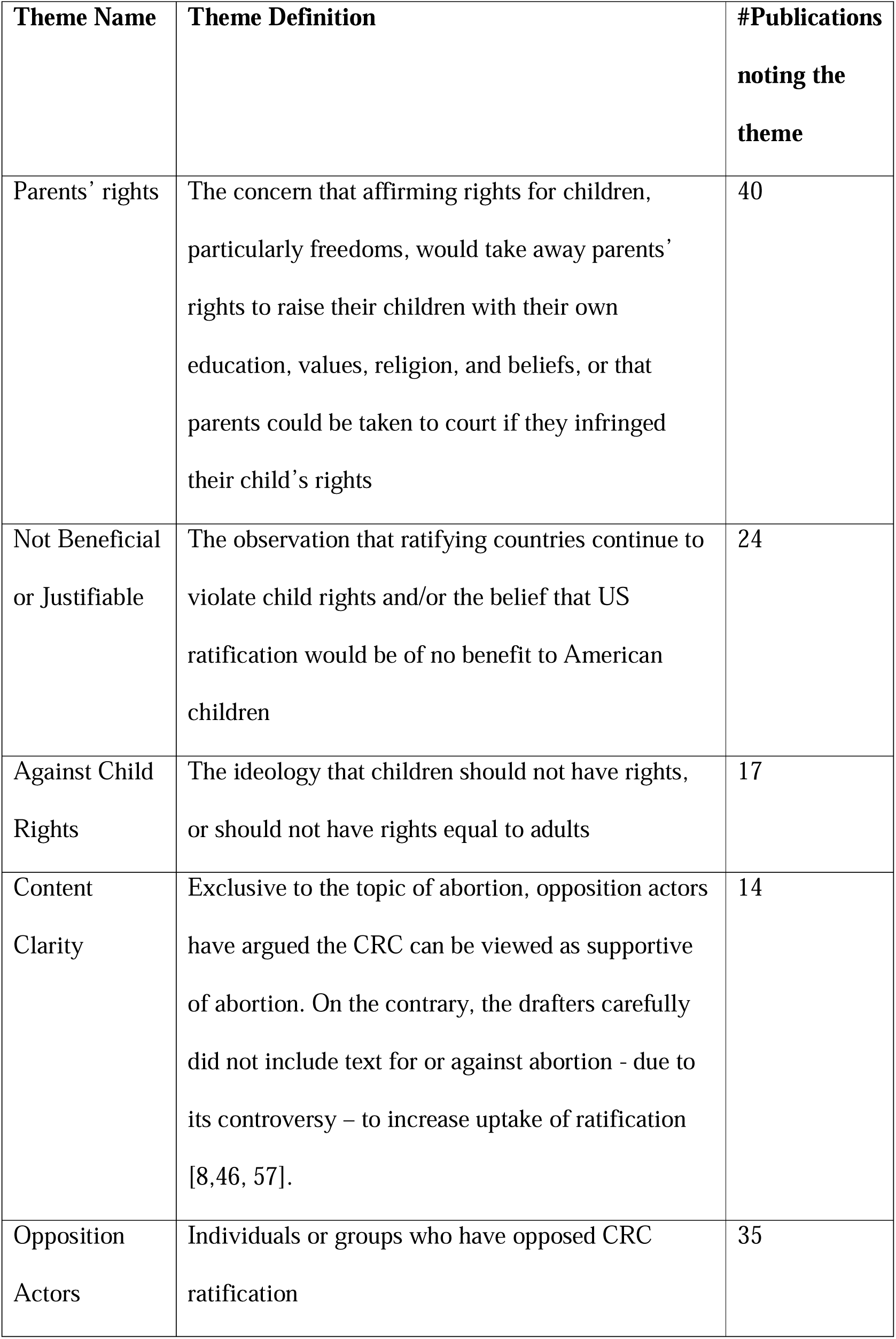
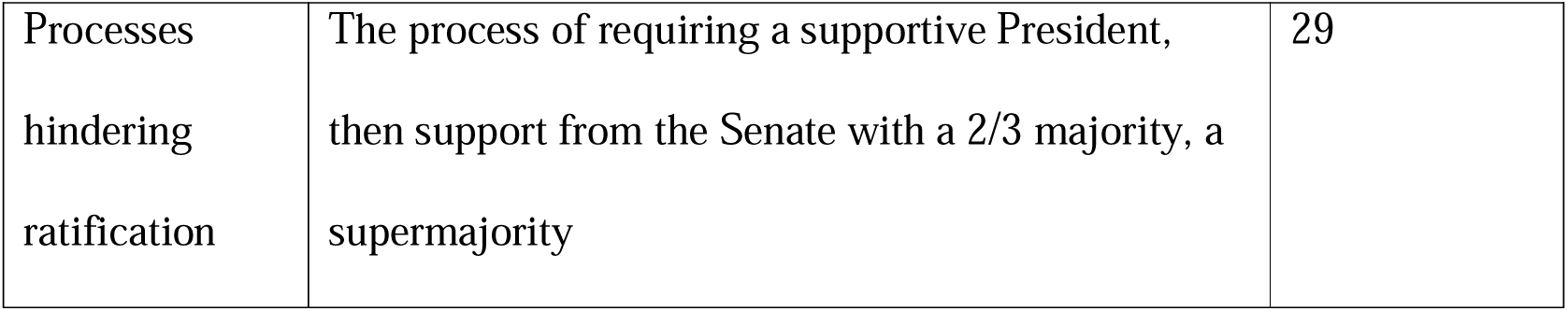
Opposition Themes Specific to CRC.

| **Theme Name** | **Theme Definition** | **#Publications**<br>**noting the**<br>**theme** |
| --- | --- | --- |
| Parents' rights | The concern that affirming rights for children, particularly freedoms, would take away parents' rights to raise their children with their own education, values, religion, and beliefs, or that parents could be taken to court if they infringed their child's rights | 40 |
| Not Beneficial<br>or Justifiable | The observation that ratifying countries continue to violate child rights and/or the belief that US ratification would be of no benefit to American children | 24 |
| Against Child<br>Rights | The ideology that children should not have rights, or should not have rights equal to adults | 17 |
| Content<br>Clarity | Exclusive to the topic of abortion, opposition actors have argued the CRC can be viewed as supportive of abortion. On the contrary, the drafters carefully did not include text for or against abortion - due to its controversy – to increase uptake of ratification [8,46, 57]. | 14 |
| Opposition<br>Actors | Individuals or groups who have opposed CRC ratification | 35 |
| Processes<br>hindering<br>ratification | The process of requiring a supportive President,<br>then support from the Senate with a 2/3 majority, a<br>supermajority | 29 |

During the thematic analysis it became clear most of the publications included multiple themes on reasons why the US <u>should</u> ratify the CRC. The ‘Pro-ratification’ themes in the second column will not be discussed further, because pro-ratification themes do not answer the research question why the CRC has not been ratified. Multiple publications from academics, professional groups, and civil society organizations include pro-ratification themes [37–46]. Likewise, ‘content’ opposition themes will not be discussed, because the content of the CRC was found to be satisfactory enough for all other UN member states to ratify it.

### Opposition Themes

Themes arising during the literature analysis became recurrent during the open coding, regardless of the discipline or background of the source. Initially, each opposition argument was coded as a separate theme, but with continued reading and coding it became clear several opposition themes were not only repeated in the CRC literature but have been opposition arguments against international human rights treaties in America for decades. In fact, many were found to be detailed in a typology of objections to ratification in Natalie Kaufman’s book, *Human Rights Treaties and the Senate: A History of Opposition*, which was referenced by multiple publications included in this study [47]. Therefore, these arguments from the opposition were collected into a super-theme titled “Historical Opposition to International Treaties” for several reasons. First, they are almost exclusively context-related opposition.

Second, enmity towards international human rights treaties from the US has existed since the 1950s, beginning with American opposition to the Genocide Convention, and for which generations of political and legal scholars have provided valid counterarguments that need not be repeated here [40,47–50]. Finally, focussing on these arguments does not advance scholarship on explaining reasons why the CRC has not been ratified. The ‘Historical Opposition to International Treaties’ themes are listed in order of theme frequency in Table 3, with definitions.

### Opposition Themes Specific to CRC

The remainder of the opposition themes are content- and context-specific arguments put forth by opposition actors against the CRC specifically, included in Table 2 and defined in Table 4. Again, they will not be discussed, because they are primarily arguments against, not reasons why the non-ratification of the CRC, and Convention scholars have already refuted them elsewhere [46,52–56].

### Opposition Actors and Processes

The themes of opposition actors and processes must be combined because the two are characteristically intertwined regarding CRC ratification. The most critical finding within the literature is that the treaty ratification process in the US depends heavily on the action of individuals. For example, President George Bush Sr. did not sign the Convention when it was first adopted by the UN, despite it having already received the approval of the Senate and the House, both of which had passed Resolutions with large majorities sponsored by both sides urging Bush to sign it and submit it to seek the advice and consent of the Senate to its ratification [58–60]. Likewise, Clinton could have submitted it after it was signed by his administration, but he did not, despite a bipartisan Senate bill urging him to sign and submit [7, 61]. Later, the Chair of the Foreign Relations Committee, Republican Senator Jesse Helms, opposed the CRC after it was signed by the Clinton administration, indicating, “as long as I am chairman of the Senate Committee on Foreign Relations, it is going to be very difficult for this treaty even to be given a hearing” [62]. Farther back, treaty scholars argue that it was Republican Senator Bricker’s proposed Bricker Amendment to the Constitution, and his resistance to the Genocide Convention backed by conservative lobby in the 1950s which has been instrumental in setting a precedent which has created the American legacy of non-ratification of human rights treaties [47,48,63]. Most recently, the Obama administration expressed support for the CRC, but faced a Senate Resolution put forth by Republican Senator Jim DeMint, with 37 Republican Senator co-sponsors – more than the number needed to veto – resolving that “the President should not transmit the Convention to the Senate for its advice and consent” [64]. In fact, the civil society lobbyists opposed to the CRC can be traced to eight conservative Christian groups and distilled further to a handful of conservative Christian leaders, who worked to influence decisions in the Senate on this and other human rights treaties (Opposition Actors in Table 2) [7,37,53,65–67]. The process of ratification of the CRC highlights how an extraordinarily small number of actors can influence major policy decisions.

### Stage 2: Interviews

Invitations for interview were extended to experts working in the fields of global health, human rights, child rights, US law, and political lobbyists. On reflection of the inherent bias of interviewing only pro-ratification informants, invitations were also extended to the eight lobby organizations who were known to oppose the CRC at the time of the original adoption. Only one opposition informant kindly agreed to be interviewed. Supplement 4 summarizes the potential informants to whom an invitation for interview was extended.

Ultimately, 14 informants were interviewed by transcribed Teams video call between July 19 and September 6, 2023, lasting 30-90 minutes each. The interviews were evaluated using the COREQ consolidated criteria for reporting qualitative research 32-item checklist for interviews and focus groups [68]. Table 5 summarizes the informants, their job description, type of organization, and area of expertise.

**Table 5:**
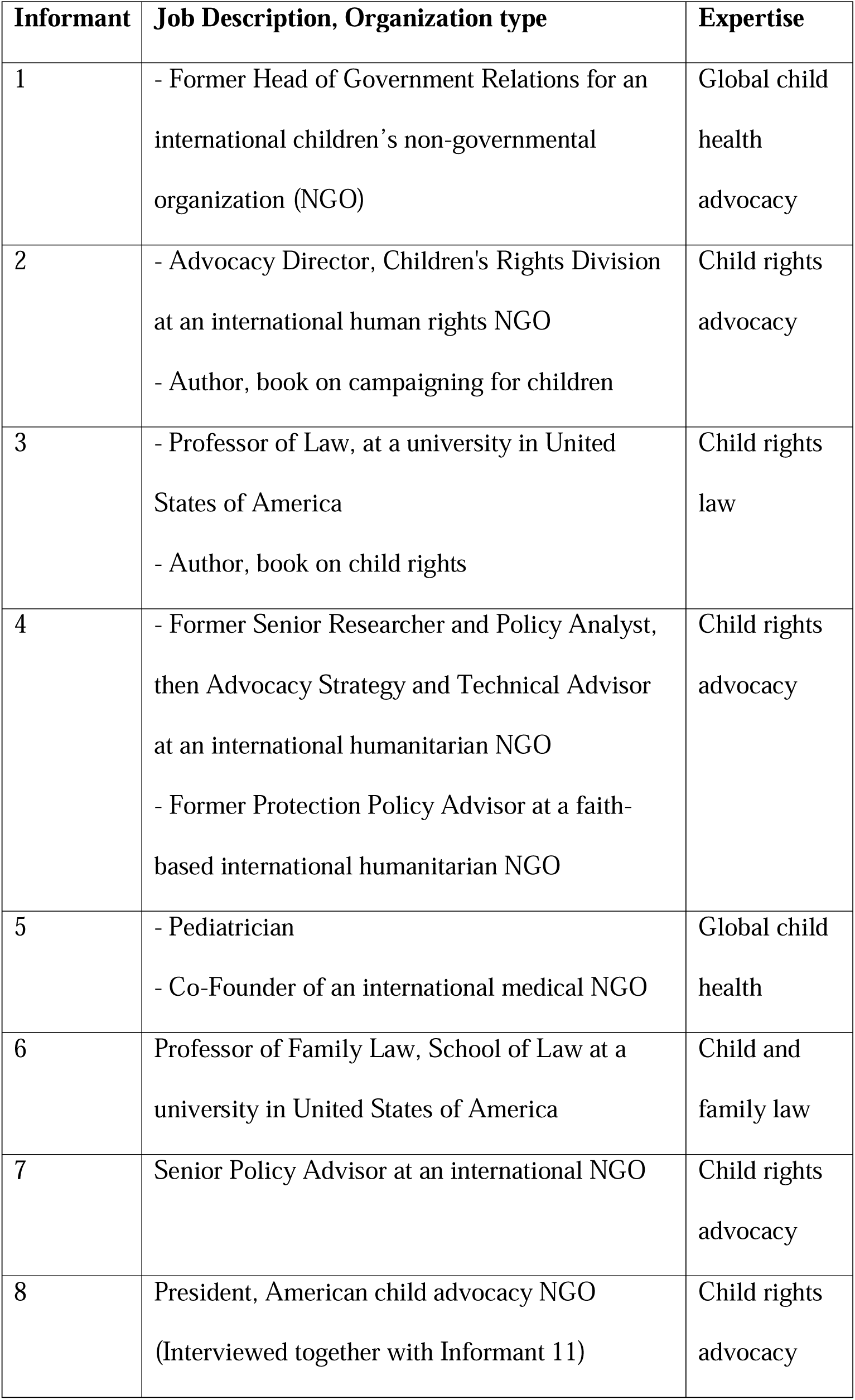
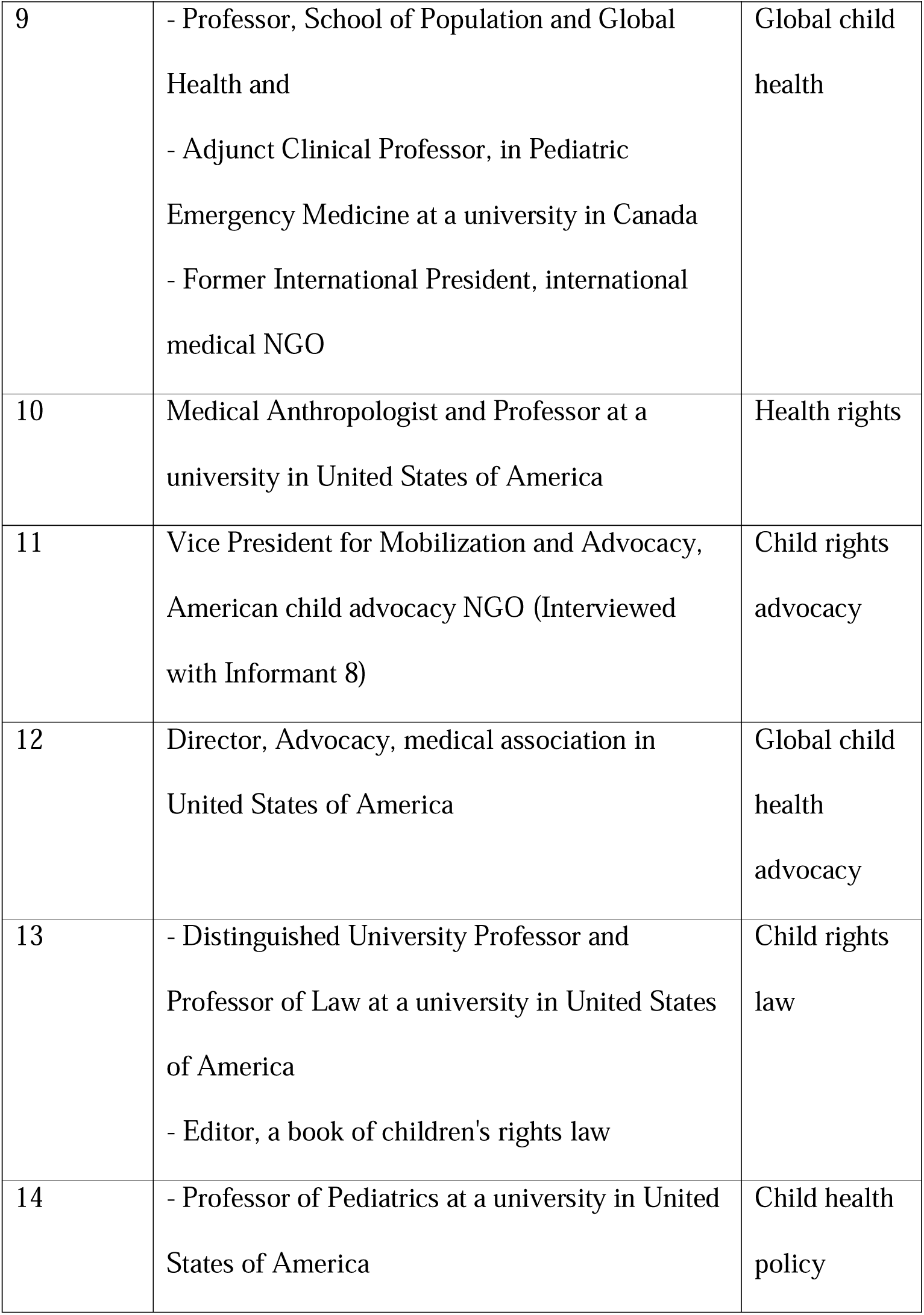
Informant Details.

| **Informant** | **Job Description, Organization type** | **Expertise** |
| --- | --- | --- |
| 1 | - Former Head of Government Relations for an international children's non-governmental organization (NGO) | Global child health advocacy |
| 2 | - Advocacy Director, Children's Rights Division at an international human rights NGO<br>- Author, book on campaigning for children | Child rights advocacy |
| 3 | - Professor of Law, at a university in United States of America<br>- Author, book on child rights | Child rights law |
| 4 | - Former Senior Researcher and Policy Analyst, then Advocacy Strategy and Technical Advisor at an international humanitarian NGO<br>- Former Protection Policy Advisor at a faith-based international humanitarian NGO | Child rights advocacy |
| 5 | - Pediatrician<br>- Co-Founder of an international medical NGO | Global child health |
| 6 | Professor of Family Law, School of Law at a university in United States of America | Child and family law |
| 7 | Senior Policy Advisor at an international NGO | Child rights advocacy |
| 8 | President, American child advocacy NGO (Interviewed together with Informant 11) | Child rights advocacy |
| 9 | - Professor, School of Population and Global Health and<br>- Adjunct Clinical Professor, in Pediatric Emergency Medicine at a university in Canada<br>- Former International President, international medical NGO | Global child health |
| 10 | Medical Anthropologist and Professor at a university in United States of America | Health rights |
| 11 | Vice President for Mobilization and Advocacy, American child advocacy NGO (Interviewed with Informant 8) | Child rights advocacy |
| 12 | Director, Advocacy, medical association in United States of America | Global child health advocacy |
| 13 | - Distinguished University Professor and Professor of Law at a university in United States of America<br>- Editor, a book of children's rights law | Child rights law |
| 14 | - Professor of Pediatrics at a university in United States of America | Child health policy |

### Generation of Themes from Interviews

Informants provided paradoxical data. On the one hand, there were no opposition arguments for which they did not have an easy counterargument. For example, on the question of sovereignty, Informant 13 observed, “the US ratified the International Covenant on Civil and Political Rights and sovereignty didn’t crumble, so it’s possible”, and Informant 2 highlights that the US did ratify the two optional protocols, so already the US must report to the UN Committee on the Rights of the Child. Regarding the Legal theme, Informant 14 notes, “lawyers, our laws, are critical for a lot of things, but how we raise our families, how communities help families are based on norms and the way I think about it is these norms are not, cannot be mandated, they must be felt”. Informant 7 commented that to accept American exceptionalism as an argument for not ratifying the CRC, “I think would logically have to accept, then, that there’s no further improvements the US can make, and I would disagree with that”. Informant 8 reprehends the opposition actor’s prioritization of banning books; meanwhile American children are increasingly dying from gun violence. Informants 5 and 11 both observe the insincerity of the “family values” and the parents’ rights arguments, because the opposition actors who promote these arguments do not champion refugee families at the border, or parents who support their LGBTQ+ children. In response to the idea that children should not have rights, Informant 3 emphasizes “those are the arguments that have been used to deprive women of rights, that were used in the United States to deprive Black people of rights, that have given second class citizenship to Chinese American rail workers… those are the arguments that are the history of oppression in the United States”, and that, “the key here is it is not a zero-sum game; when you give children rights, it does not detract from other people’s rights”. Similarly, Informant 10 observes, “children are human, and humans all have a right to basic needs and safety and security and food and healthcare.” Informant 1 reflects that the American public endorses the CRC; “[t]here’s been a lot of polling done amongst Republican and Democratic voters, so I think it is absolutely, in my opinion, it does represent the American values.”

On the other side of the paradox, the informants have lost all expectation of ratification, made clear in the language they use on the likelihood of ratification; “there is not the political will”, “dead in the water”, “unlikely that it is going to fly”, “political suicide”, “harder and harder”, “killing it permanently”, “such a monumental lift”, “politically catastrophic”, “that’s magical thinking”, “the Convention would take a lot of political capital to move”, “I’m very sceptical”, “I do not think the gerontocracy right now cares”, and “the atomic bomb of topics that no one’s going to touch”. As can be seen, there is a contradiction of pro-ratification informants providing easy counterarguments to opposition, while remaining doubtful on the possibility of ratification. Of the pro-ratification informants, only one remains optimistic that the CRC will be ratified.

Importantly, data from all the informants confirmed the validity of the data from the literature on historical opposition themes. However, new themes were revealed during the interviews, which were not apparent in the literature. They are summarized as Chilling Effects, and Children Cannot Vote. The emergent themes are listed and defined in Table 6.

**Table 6:** Emergent Opposition Themes from Interviews.

|  | **Theme Name** | **Theme Definition** | **#Informants noting the theme** |
| --- | --- | --- | --- |
| Chilling Effects<br>(Effects of child rights advocates abandoning lobby efforts to have the CRC ratified) | Off the Agenda | Ratification is not a current issue in Congress; politicians are not interested | 12 |
|  | Too Difficult to Ratify | Resignation to the idea that due to historical opposition and the US international policy norm of not ratifying international human rights treaties, the US will likely never ratify the CRC | 11 |
|  | Other Efforts | Would-be CRC supporters have turned to other efforts to improve the rights and lives of children | 9 |
|  | Silent Supporters | Actors who support child rights and/or ratification of the CRC, who are not actively seeking to have it ratified | 8 |
| Children Cannot Vote |  | Unlike other human rights' movements, the beneficiaries of the CRC do not have electoral agency so politicians do not prioritize their needs | 6 |

### Effects of Abandoning Lobby Efforts to Ratify the CRC

All of the pro-ratification informants endorse the ideals of the CRC and believe it should be ratified in principle, however, most perceive it would require insurmountable effort to have it ratified. The pro-ratification informants view getting the CRC passed by the US Congress as a virtual impossibility, observing that the CRC is off the agenda, too much work to ratify, and/or that other endeavours would be more fruitful in affirming the rights and improving the lives of children (Tables 2&6).

### Children Cannot Vote

It was the direct or indirect observation of all of the pro-ratification informants that politicians work to remain in office; therefore, they do not prioritize children’s issues because children cannot cast ballots. Informant 9 noted this theme most succinctly, observing, “Children are the constituents of nobody; they don’t vote, the same thing as migrants.” While it is important to acknowledge that other groups like women, and Black, Indigenous and People of Colour have not yet realized full equality in America, children do not even have the voice afforded to them. Informant 2 observed that the Parkland students protesting for gun reforms could only threaten that one day they would have a vote.

## Discussion

This study aimed to determine why the USA has not ratified the UN Convention on the Rights of the Child. The literature was found to be wanting on primary research which has investigated the question, so a mixed methods approach was designed to answer the question. The expert interviews were critical to answering the question, because the informants identified both the chronicity of the veto, and the pro-ratification actors giving up hope as factors in the non-ratification.

### Walt and Gilson’s Health Policy Triangle

Reviewing the non-ratification of the CRC using the framework of the health policy triangle, it is evident that pro-ratification *actors* within America far outweigh opposing actors, and the *context* and *content* of the CRC also favour ratification – the literature weighs heavily on pro-ratification themes, including offering feasible solutions to opposition arguments - ultimately, it is the *process* and *key actors* which have prohibited ratification. As outlined in Table 1, a veto from the Senate requires not a majority, but opposition votes of only one-third plus one senators for it to fail, or even by leaving it off the agenda [11]. Compared to other countries, the American system of “a two-thirds majority is a more stringent requirement than that of any other nation for ratifying treaties”, making ratification extremely challenging, as the informants observed [53]. The USA has ratified only three of nine of the core UN conventions, and only five of eighteen treaties if optional protocols are included; America has set a precedent of not ratifying multiple prior human rights treaties [69]. Thus, the process of CRC non-ratification by the United States began far earlier than the drafting of the CRC; it was set up to fail.

### Tsebelis’ Veto Player Theory

Further, this research has found that actors are intimately tied to process, and certain actors - veto players - have been influential in the non-ratification. Tsebelis’ veto player theory describes how the greater number of veto players in a political system increases the chance of veto of a particular policy, and a tendency to the status quo [24]. Ratification of human rights treaties in America relies on assent from both the President and the Senate, two veto points (Table 1). Critically though, this research has found that the CRC has been chronically vetoed, and the longer it is vetoed, the more likely ratification is chilled, and the status quo remains.

### A New Policy Analysis Framework – The Veto Fulcrum

This research therefore proposes that veto players are the fulcrum of a balance. By adapting Walt and Gilson’s health policy triangle with the veto actors at the centre of the triangle, content/context as the weights, and process/lobby actors as the moveability of the fulcrum, it becomes evident that ratification of an international treaty in the American system depends primarily on the decisions of individuals. Put simply, the relative weights of arguments for or against ratification are not of prime relevance, but rather, ratification/change depends on the action of the player in the veto position, and the motility of the fulcrum, the process. Figure 3 illustrates the ratification of a policy for which context and content favour ratification, and how process allows single veto players to impede ratification.

**Fig 3:**
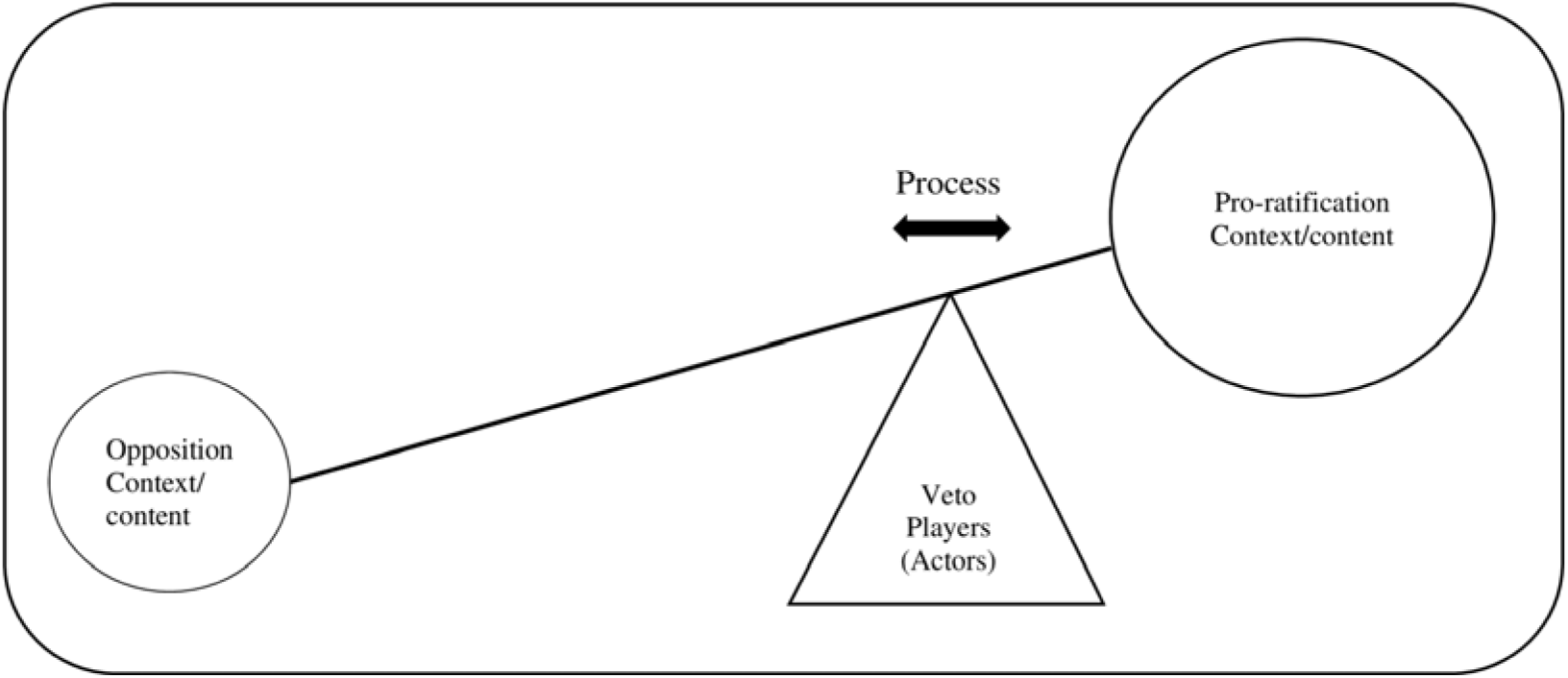
How Veto Players and Process Obstruct Ratification.

The history of non-ratification of the CRC highlights how the decision of a single individual can veto a policy. For example, in the case of the CRC in America, consider the outcome if a ratification-friendly president had been in power at the time of its adoption instead of George Bush Sr., when it had already received majority approval from both the Senate and the House [58–60]. Why did Clinton refuse to submit the CRC to a welcoming Senate, after it was signed? [7, 61] If, during Obama’s time as President, the Senate had been pro-ratification, then both President and Senate would have viewed it favourably. Early in America’s history with human rights treaties, if Republican Senator Bricker had not resisted the Genocide Convention, it is plausible that all of the human rights treaties before the CRC would have been accepted as American political norms, and the CRC would have been ratified simply as one of many [48, 63].

Thus, treaty ratification is determined by the decision of the veto player(s) at the fulcrum, not by the relative weights of opposition and support, and a tipping point could theoretically occur by changing the mind of a veto player or changing the veto player. In the process of CRC ratification, it should be possible to move the position of the fulcrum by convincing the veto player(s) at the fulcrum on the value of ratification. In other words, if the current President and Senate understood the American public is pro-ratification, these players in veto positions should want to satisfy their constituents, moving the green arrow in Figure 4 [14]. On the other hand, the influence of powerful opposition lobby organizations, who are often private special interest actors, can move the fulcrum away from good health policy (red arrow Fig 4). In the case of prolonged veto, the process is affected because would-be advocates abandon hope of ratification, which cements the fulcrum in place, eliminating the green arrow in Figure 4.

**Fig 4.**
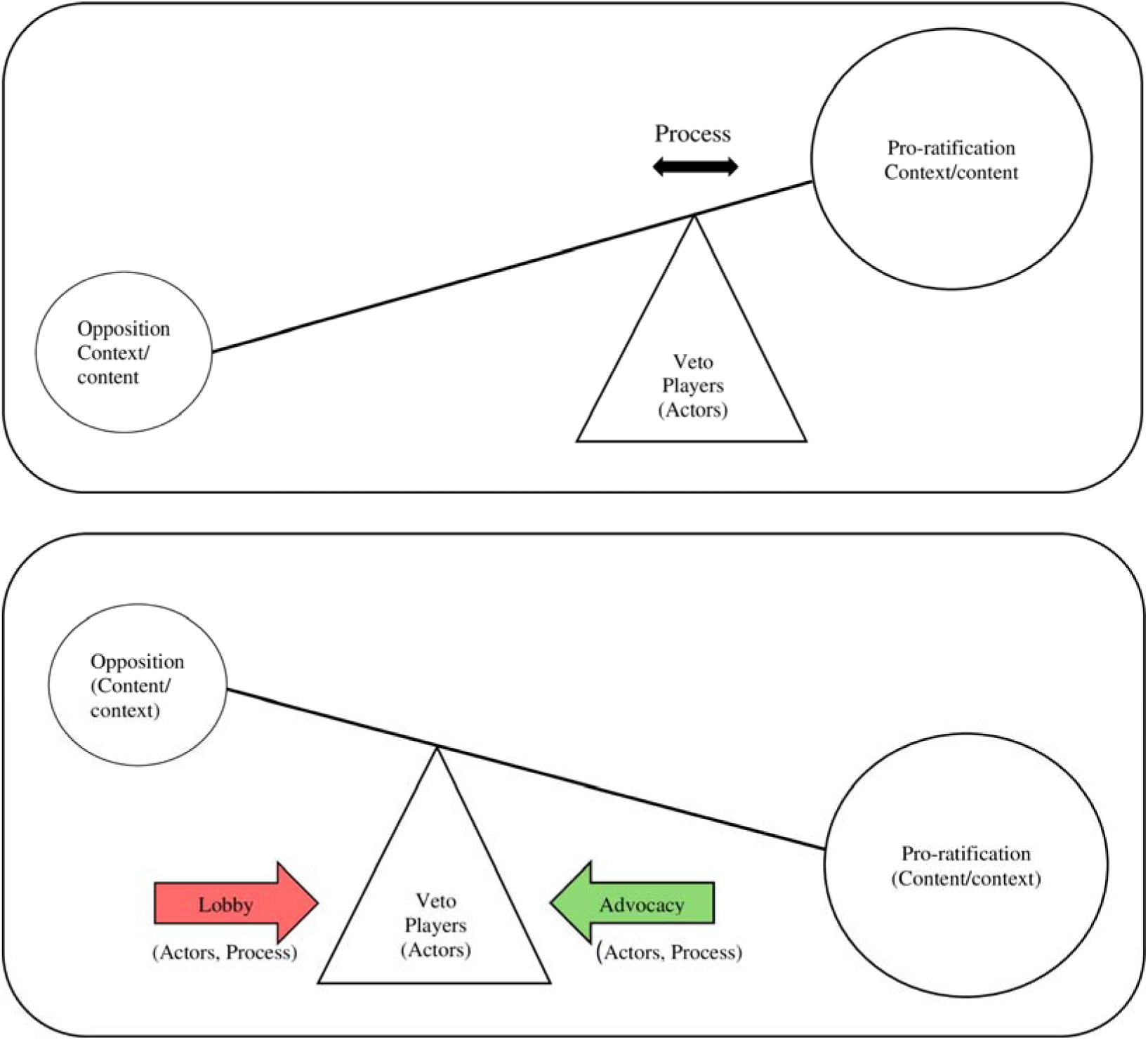
Moving the Veto Fulcrum.

The research exercise of identifying and categorizing themes of opposition led to the discovery that most of the opposition themes in the literature were *arguments against* ratification, rather than *reasons why* the US has not ratified the CRC. This distinction led to the creation of the veto fulcrum framework to answer the research question. Most of the arguments against the CRC have been general arguments against the US ratifying human rights treaties for decades yet have been easily refuted by proponents of ratification. However, the pattern of non-ratification became one of the *reasons why* the CRC is not ratified; non-ratification of human rights treaties has become the status quo in America, because veto players at key moments in history have chilled prospects for ratification, causing child rights proponents to abandon hope and turn their energy elsewhere.

### Veto Fulcrum Framework

Understanding the political power which actors can exert on and from the veto fulcrum is a framework which can be employed to understand other political systems which tip the balance away from health-friendly policies. Actors at a veto fulcrum make big decisions, often against the wishes of the population, even in otherwise democratic systems. Examining the non-ratification of the CRC also highlights the political power of lobby organizations (red arrow, Figure 4), many of whom are single-interest groups, political action committees (PACs), industry, individual wealthy donors, or private corporations. Indeed, this research is not the first to find the government of the United States does not make policy decisions based on the will of the general population; research by Gilens and Page found that US government policy decisions align more closely with the preferences of wealthy elites than with the average American [70]. The CRC has not been ratified because initially powerful opposition lobbyists influenced the actors at the veto fulcrum, and the fulcrum has subsequently become immovable, giving one actor absolute power in a system which has been the antithesis of the democratic process. Case in point: at this moment, a single actor - the President of the US - is exerting political power by leaving the CRC off the agenda, effectively vetoing it (Step 3, Table 1).

Examining the US non-ratification of the CRC exemplifies how the veto fulcrum framework can identify the obstruction to health and other good governance policy change; for example, each of the following policy arenas should be evaluated using the veto fulcrum framework (each could be its own focus of research, which is beyond the scope of this paper). For example, the Supreme Court of the US is not democratically elected yet shifted the veto fulcrum on several policy issues including abortion and LGBTQ+ rights, away from good health policy [71,72]. Importantly, the same conservative lobbyists who opposed the CRC (Table 2, and red arrow in Figure 4) have also backed the appointment of the new Supreme Court Justices, and written the Republican presidential campaign Project 2025 [53,65–67,73,74]. In another example of an undemocratic veto fulcrum in international governance, the five permanent members of the UN Security Council (UNSC - China, France, Russia, the United Kingdom, and the United States) each holds a veto vote on all Council decisions which cements the veto fulcrum of the UNSC, creating a stalemate on decisions which affect the security of the world’s people. It is urgent to examine who influences the UNSC veto players, because at the time of this writing ten of the last fifteen vetoes of the UNSC have been on the “the situation in the Middle East” [75]. In an illustration of private actors as veto players, governance of media and social media can be analyzed as veto fulcra, because they are not democratically elected, yet they determine what news is shared, to whom, and how, by vetoing the people and information which are presented on their platforms, meanwhile governments are deciding which media platforms are even legal [76]. Likewise, governance of artificial intelligence (AI) must be similarly scrutinized using the veto fulcrum, to ensure that data collected from the people which is being used to generate AI is given freely with informed consent and used in a fair and just way [77]. Finally, in the sphere of global health, the veto players who determine funding for projects in the Global South are typically donors from the Global North acting as veto players, the most dramatic example of which has been the recent withdrawal of United States Agency for International Development (USAID) funding [78].

As with the CRC, when a health or other good governance policy does not pass, or alternately, when a policy passes which contravenes health, environmental, or climate protection, it is important to start asking questions. This research proposes that the veto fulcrum is an instrument which can answer the questions. Identifying and analyzing the actors at the veto fulcrum and the influences on them is generalizable to all policies, including other democracies, and other forms of governance within state, private, and international institutions. Firstly, by examining health policies and other governance decisions using a veto fulcrum framework approach, policy analysts can identify these undemocratic processes and begin to challenge them. It is essential to ask five questions: 1) who is at the veto fulcrum, 2) how did they get there, 3) who is influencing them, 4) what is their position on the issue, and 5) can they be moved on the issue? Secondarily, to address other governance structures including undemocratic systems where veto players are appointed or self-appointed (as in private actors, dictatorships, de facto governments), identifying who is at the veto fulcrum can be a starting point in any political system to challenge the existing governance towards ensuring veto players are fairly and democratically elected, and work towards the betterment of one health.

## Conclusions

This study revealed that the process within the American political system for ratification of the CRC – and indeed, for all international human rights treaties - is obstructed by the treaty ratification process itself, allowing single actors as veto players to exert absolute decision-making power on issues which affect the lives of their constituents. The veto fulcrum is a new policy evaluation framework which can critically analyse health and other good governance policies to ensure health policies are decided fairly and in the best interest of the people, particularly the most vulnerable people.

## Data Availability

Interview transcript data is protected, as agreed by informant consent and confidentiality agreement.

## Acknowledgments

This study topic was originally a master’s thesis research project. I am grateful to the invaluable contributions of project module organizer, Alexandra Conseil, and project supervisor, Dr. Rachel Hammonds, as well as the study participants, as this manuscript would not have been possible without their time and expertise. Thanks to Dr. Shashika Bandara, whose work I have admired, and who was kind enough to provide feedback on the manuscript from a stranger who reached out on LinkedIn.

## Author Contributions

LH conceptualised the research design and proposal, collected data, undertook the analysis, and wrote the manuscript.

## Disclosure Statement

The author has no competing interests.

## Ethics and Consent

This research has received ethical approval from the London School of Hygiene and Tropical Medicine’s (LSHTM) MSc Research Ethics committee ID#28417 and qualifies for exemption under “Research Involving Human Subjects Eligible for Exemption Under 45 CFR 46.104(d)”, Chart 4 from the US Department of Health and Human Services Human Subject Regulations. Documentation from both institutions is available. Participants gave informed consent to participate in the study either as signed consent (Supplement 5), or verbally at the beginning of the recorded interview before taking part and were provided transcripts of the interview at request.

## Funding Information

None.

## Preprint

Preprint manuscript is available in medRxiv preprint here: https://www.medrxiv.org/content/10.1101/2024.09.05.24312304v2.

## Paper Context

- **Main findings** Whereas previous literature focuses on pros and cons of ratification, this study answers *why* the United States of America has not ratified the Convention on the Rights of the Child despite overwhelming support for it.
- **Added knowledge** Revising Walt and Gilson’s health policy triangle and drawing on Tsebelis’ veto player theory a new policy analysis framework approach is introduced – the veto fulcrum – which highlights the intimate correlation between actors and processes and their relative importance over content and context factors in policy decisions.
- **Global health impact for policy and action** The veto fulcrum framework is an accessible policy analysis tool which can be applied to understand policy decisions, by identifying and examining barriers to ratification or enactment of health or other good governance policies.

## Data Availability

Interview transcript data is protected, as agreed by informant consent and confidentiality agreement, however, the parts of the data transcript data which were not identified in interviews as “off the record” can be made available on reasonable request.

## Notes

### Competing Interest Statement

The authors have declared no competing interest.

### Funding Statement

This research received no specific grant from any funding agency in the public, commercial, or not-for profit sectors.

### Author Declarations

This research has received ethical approval from the London School of Hygiene and Tropical Medicine (LSHTM) MSc Research Ethics committee ID#28417 and qualifies for exemption under Research Involving Human Subjects Eligible for Exemption Under 45 CFR 46.104(d), Chart 4 from the US Department of Health and Human Services Human Subject Regulations. Documentation from both institutions is available. Participants gave informed consent to participate in the study either as signed consent, or verbally at the beginning of the recorded interview before taking part, and all participants provided written consent for their names, professions, and quoted material to be published.

### Summary of Updates

Manuscript has been accepted for publication by Global Health Action, but only if informant data is anonymized.

