## Supplement1to5 for "Why has the United States of America not ratified the United Nations Convention on the Rights of the Child? The veto fulcrum as a new health policy analysis framework"

**Supplemental Material Checklist**

1. Interview Outline Questions p.2
2. CASP Checklist p.3
3. Publications and CASP Appraisals p.4
4. Informant Selection p.12
5. Informed Consent Form p.13

**Supplement 1: Interview Outline Questions**

Version 1.2; July 18, 2023

1. How would you like me to present your name and educational background?
2. What is your job title, job description, area of expertise?
3. Do you think it is important for the US to ratify the UNCRC?
4. Why or why not?
5. What, in your opinion, are good reasons for US ratification?
6. What, in your opinion, are reasons against US ratification?
7. Do you believe global child health and health care can be influenced by US ratification?
8. Why or why not?
9. To your knowledge, have there been efforts to motivate the US to ratify the UNCRC?
10. What were the efforts?
11. What were the successes/failures of the efforts?
12. What are the influential powers working towards and against the ratification?
13. Are they within or outside the US?
14. How has power affected the non-ratification status of the UNCRC?
15. What power?
16. Are/were the influences overt or covert? Describe
17. What do you imagine would be effective means to convince the US to ratify? (I will give examples such as, but not limited to civil society lobbies, pressure from other UN states, citizen movements, financial backing of parties, pressure from international organizations, corporate involvement etc)
18. What do you imagine would “backfire” as efforts to convince the US to ratify? (same examples)
19. Do you believe there are other activities which would improve child health and human rights more than US ratification?
20. What are they?
21. Please share any other information you feel is useful and we have not discussed.
22. Do you think a children’s movement within the USA would be an effective tool to get it ratified?
23. Why or why not?
24. What has been the role of the following themes in non-ratification?
    a. Sovereignty
    b. Federalism
    c. Parent rights
    d. Need to change the laws
    e. American constitution already protects children adequately
    f. Children should not have rights (esp participation rights)
    g. American exceptionalism
    h. It is not beneficial (example, child rights are violated in other countries, time and effort is better spent on other advocacy, ratification would not improve rights, no certainty it will help/not harm, no one cares about child rights)
    i. In the queue behind CEDAW
    j. Off the agenda
    k. Opposition actors (conservative right)
    l. US distrust of UN
    m. No enforcement mechanisms
25. Please refer me to anyone who works in the field of child health, child rights, human rights, and/or global health, whom you think would have some insight into my questions.

**Supplement 2: CASP Appraisal Checklist**

All 68 papers included in the study were subjected to critical appraisal, with these six questions in mind:

1. Where did you find the information?
2. How has the author/ speaker come to their conclusions?
3. When was this written/said?
4. What is it and what are the key messages or results/findings?
5. Who has written/said this?
6. Why has this been written/ said?

On the basis of these six questions, the papers were classified as follows:

A. High quality peer-reviewed study, with no major flaws in design
B. High quality study that is not peer-reviewed. Few concerns with using its findings
C. Medium quality study – which may or may not be peer-reviewed. Usable, but with some small concerns
D. Medium quality study. Usable, but with caution
E. Low quality study. Findings not deemed reliable enough to use

**Supplement 3: Publications Included in the Study and CASP Appraisal**

| CASP  Grade | Publication | Database  Source | Pro-/  Neutral/  Opposed | #Themes  Identified | Discipline |
| --- | --- | --- | --- | --- | --- |
| A | Wegman ME. Foreign Aid, International Organizations, and the World’s Children. Pediatrics. 1999 Mar 1;103(3):646–54. | Medline | Pro | 7 | Medicine/ Public Health |
| A | Tait CA, Parnia A, Zewge-Abubaker N, Wong WH, Smith-Cannoy H, Siddiqi A. Did the UN convention on the rights of the child reduce child mortality around the world? An interrupted time series analysis. BMC Public Health. 2020 May 18;20:707. | Web of Science | Pro | 4 | Medicine/  Public Health |
| A | Clark H, Coll-Seck AM, Banerjee A, Peterson S, Dalglish SL, Ameratunga S, et al. A future for the world’s children? A WHO–UNICEF–Lancet Commission. The Lancet. 2020 Feb 22;395(10224):605–58. | Web of Science | Neutral | 5 | Medicine/ Public Health |
| A | Hoffman SJ, Baral P, Rogers Van Katwyk S, Sritharan L, Hughsam M, Randhawa H, et al. International treaties have mostly failed to produce their intended effects. Proc Natl Acad Sci. 2022 Aug 9;119(32):e2122854119. | Grey Literature | Opposed | 2 | Medicine/ Public Health |
| A | Comstock AL. Legislative veto players and human rights treaty signature timing. J Legis Stud. 2022 Sep 20;1–29. | Google Scholar | Neutral | 8 | Political Science |
| A | Moravcsik A. Chapter 6. The Paradox of U.S. Human Rights Policy. In: Chapter 6 The Paradox of US Human Rights Policy [Internet]. Princeton University Press; 2009 [cited 2023 Sep 18]. p. 147–97. | Grey Literature | Neutral | 18 | Political Science |
| A | Palmer A, Tomkinson J, Phung C, Ford N, Joffres M, Fernandes KA, et al. Does ratification of human-rights treaties have effects on population health? The Lancet. 2009 Jun 6;373(9679):1987–92. | Grey Literature | Opposed | 3 | Medicine/ Public Health |
| B | Ruck MD, Keating DP, Saewyc EM, Earls F, Ben-Arieh A. The United Nations Convention on the Rights of the Child: Its Relevance for Adolescents. J Res Adolesc. 2016;26(1):16–29. | Web of Science | Pro | 11 | Psychology |
| B | Haggerty RJ. The Convention on the Rights of the Child: It’s Time for the United States to Ratify. Pediatrics. 1994 Nov 1;94(5):746–7. | Grey Literature | Pro | 3 | Medicine/ Public Health |
| B | Minow M. What Ever Happened to Children’s Rights. Minn Law Rev [Internet]. 1995 Jan 1; Available from: <https://scholarship.law.umn.edu/mlr/1897> | Grey Literature | Pro | 11 | Law |
| B | Scherrer J. The United Nations Convention on the Rights of the Child as Policy and Strategy for Social Work Action in Child Welfare in the United States. Soc Work. 2012 Jan;57(1):11–22. | Medline | Pro | 15 | Social Work |
| B | Browning DS, Witte J Jr. Christianity’s Mixed Contributions to Children’s Rights. Zygon®. 2011;46(3):713–32. | Web of Science | Pro | 14 | Religious Studies |
| B | Yanghee Lee. The USA and non-ratification of the convention on the rights of the child. Child Welfare. 2010 Oct 9;89(5):15–9. | Grey Literature | Pro | 2 | Rights Activist |
| B | The `Punitive Turn’ in Juvenile Justice: Cultures of Control and Rights Compliance in Western Europe and the USA - John Muncie, 2008 [Internet]. [cited 2023 Jul 5]. Available from: <https://journals-sagepub-com.ez.lshtm.ac.uk/doi/abs/10.1177/1473225408091372> | Scopus | Pro | 9 | Law |
| B | Todres J. Children’s health in the United States: assessing the potential impact of the convention on the rights of the child. Child Welfare. 2010 Oct 9;89(5):37–56. | Medline | Pro | 11 | Law |
| B | Gardinier M. Introduction: why should the united states ratify the convention on the rights of the child? Child Welfare. 2010 Sep 1;89(5):7–14. | Grey  Literature | Pro | 11 | Rights Activist |
| B | Blanchfield L. The United Nations Convention on the Rights of the Child. U N Conv Rights Child. April 1, 2013 | Grey Literature | Neutral | 23 | Political Science |
| B | Cook JT, Brown JL. Children’s rights to adequate nutritious foods in the two Americas. Food Policy. 1996 Mar 1;21(1):11–6 | Scopus | Pro | 9 | Ethics |
| B | BARTHOLET E. Ratification by the United States of the Convention on the Rights of the Child: Pros and Cons from a Child’s Rights Perspective. Ann Am Acad Pol Soc Sci. 2011;633:80–101. | Grey Literature | Pro | 20 | Law |
| B | Sen. DeMint J [R S. Text - S.Res.99 - 112th Congress (2011-2012): [Internet]. 2011 [cited 2023 Jun 23]. Available from: <http://www.congress.gov/bill/112th-congress/senate-resolution/99/text> | Grey Literature | Opposed | 10 | Political Science |
| B | Law KA. Hope for the Future: Overcoming Jurisdictional Concerns to Achieve United States Ratification of the Convention on the Rights of the Child. Fordham Law Rev. 1993 1994;62:1851. | Grey Literature | Pro | 12 | Law |
| B | Lopez, D. (2018). The time is now to ratify the convention on the rights of the child. University of San Francisco Law Review, 52(3), 477-500. | Grey Literature | Pro | 17 | Law |
| B | Guggenheim, M. (2006). Ratify the U.N. Convention on the Rights of the Child, but don't expect any miracles. Emory International Law Review, 20(1), 43-68. | Grey Literature | Pro | 12 | Law |
| B | Levy SR, Migacheva K, Ramírez L, Okorodudu C, Cook H, Araujo-Soares V, et al. A human rights based approach to the global children’s rights crisis: A call to action. J Soc Issues. 2022;78(4):1085–97. | Web of Science | Pro | 3 | Psychology |
| B | Todres J. Emerging Limitations on the Rights of the Child: The U.N. Convention on the Rights of the Child and Its Early Case Law. Columbia Hum Rights Law Rev. 1998 1999;30:159. | Google Scholar | Pro | 14 | Law |
| B | Mason MA. The U.S. and the International Children’s Rights Crusade: Leader or Laggard? J Soc Hist. 2005;38(4):955–63. | Grey Literature | Pro | 14 | Law |
| B | Stentzel LLI. Prospects for United States Ratification of the Convention on the Rights of the Child. Wash Lee Law Rev. 1991;48:1285. | Google Scholar | Pro | 22 | Law |
| B | Calciano EM. United Nations Convention on the Rights of the Child: Will It Help Children in the United States. Hastings Int Comp Law Rev. 1991 1992;15:515. | Google Scholar | Pro | 17 | Law |
| B | Henkin L. U.S. Ratification of Human Rights Conventions: The Ghost of Senator Bricker. Am J Int Law. 1995;89(2):341–50. | Grey Literature | Pro | 13 | Law |
| B | Earls F. Children: From Rights to Citizenship. Ann Am Acad Pol Soc Sci. 2011 Jan 1;633(1):6–16. | Web of Science | Pro | 20 | Law |
| B | Rutkow L, Lozman JT. Suffer the Children: A Call for United States Ratification of the United Nations Convention on the Rights of the Child. Harv Hum Rights J. 2006;19:161. | Google Scholar | Pro | 22 | Law |
| B | Engman M. And Then There Were Two: Why Is the United States One of Only Two Countries in the World That Has Not Ratified the Convention on the Rights of the Child. DePaul Int Hum Rights Law J. 2015;1:[i]. | Google Scholar | Pro | 17 | Rights Activist |
| B | Kasper J. The relevance of U.S. Ratification of the Convention on the Rights of the Child for child health: a matter of equity and social justice. Child Welfare. 2010 Oct 9;89(5):21–36. | Medline | Pro | 9 | Medicine/ Public Health |
| B | Helman C. In Favor of United States Ratification of the Convention on the Rights of the Child. Child Leg Rights J. 2019;39:191. | Google Scholar | Pro | 8 | Law |
| B | Galvin C. Opposing Viewpoints: The U.S. Should Not Ratify the United Nations Convention on the Rights of the Child. Child Leg Rights J. 2020;39. | Grey Literature | Neutral | 14 | Law |
| B | Adams C, Rubel J. Compliance issues raised by the United States’ ratification and implementation of the education articles of the convention on the rights of the child. Child Welfare. 2010 Oct 9;89(5):73–90. | Medline | Pro | 12 | Law |
| B | Kilbourne S. U.S. Failure to Ratify the U.N. Convention on the Rights of the Child: Playing Politics with Children’s Rights. Transnatl Law Contemp Probl. 1996;6:437. | Grey Literature | Pro | 23 | Law |
| B | Pizzigati K. Companion piece: the education landscape and the convention on the rights of the child. Child Welfare. 2010 Oct 9;89(5):91–102. | Medline | Pro | 16 | Political Science |
| B | Anthropology S for M. The Rights of Children. Med Anthropol Q. 2007;21(2):234–8. | Scopus | Pro | 12 | Anthropology |
| B | Nafziger JAR. State Collaboration in United States Ratification of Human Rights Treaties. ISLA J Int Comp Law. 1996 1997;3:621. | Web of Science | Pro | 8 | Law |
| B | Molnar BE. Juveniles and Psychiatric Institutionalization: Toward Better Due Process and Treatment Review in the United States. Health Hum Rights. 1997;2(2):98–116. | Medline | Pro | 6 | Medicine/ Public Health |
| B | Svevo-Cianci K, Velazquez SC. Companion piece: convention on the rights of the child special protection measures: overview of implications and value for children in the united states. Child Welfare. 2010 Oct 9;89(5):139–57. | Medline | Pro | 11 | Rights Activists |
| B | Renteln AD. Who’s Afraid of the CRC: Objections to the Convention on the Rights of the Child. ISLA J Int Comp Law. 1996 1997;3:629. | Grey Literature | Pro | 4 | Political Science |
| B | Weber MA. Global Human Rights: Multilateral Bodies & U.S. Participation. Congressional Research Service. November 23, 2018. | Grey Literature | Neutral | 9 | Political Science |
| B | Quigley J. U.S. Ratification of the Convention on the Rights of the Child. St Louis Univ Public Law Rev. 2003;22:401. | Google Scholar | Pro | 13 | Law |
| B | Nauck BJ. Implications of the United States Ratification of the United Nations Convention on the Rights of the Child: Civil Rights, the Constitution and the Family. Clevel State Law Rev. 1994;42:675. | Google Scholar | Neutral | 14 | Law |
| B | Donnolo P, Azzarelli KK. Ignoring the Human Rights of Children: A Prespective on America’s Failure to Ratify the United Nations Convention on the Rights of the Child. J Law Policy. 1996 1997;5:203. | Google Scholar | Pro | 14 | Law |
| B | Blanchfield L. The United Nations Convention on the Rights of the Child. U N Conv Rights Child. Congressional Research Service July 27, 2015 | Grey Literature | Neutral | 18 | Political Science |
| B | Uchitel J, Alden E, Bhutta ZA, Goldhagen J, Narayan AP, Raman S, et al. The Rights of Children for Optimal Development and Nurturing Care. Pediatrics. 2019 Dec 1;144(6):e20190487. | Medline | Pro | 13 | Medicine/ Public Health |
| B | Wagner JT. Child Rights in the United States: Dilemmas and Questions. In: Višnjić-Jevtić A, Sadownik AR, Engdahl I, editors. Young Children in the World and Their Rights: Thirty Years with the United Nations Convention on the Rights of the Child [Internet]. Cham: Springer International Publishing; 2021 [cited 2023 Apr 11]. p. 31–41. (International Perspectives on Early Childhood Education and Development). | Grey Literature | Pro | 23 | Education |
| B | Weissbrodt D. Prospects for Ratification of the Convention on the Rights of the Child. Emory Int Law Rev. 2006;20:209. | Google Scholar | Pro | 23 | Law |
| B | Gautam KC. Time for USA to ratify the child rights convention. Child Welfare. 2010 Oct 9;89(5):221–4. | Grey Literature | Pro | 13 | Rights Activists |
| B | Aber JL, Hammond AS, Thompson SM. U.S. Ratification of the CRC and reducing child poverty: can we get there from here? Child Welfare. 2010 Oct 9;89(5):159–75. | Grey Literature | Pro | 12 | Psychology |
| B | Lichtsinn H, Goldhagen J. Why the USA should ratify the UN Convention on the Rights of the Child. BMJ Paediatr Open. 2023 Feb 1;7(1):e001355. | Medline | Pro | 15 | Medicine/ Public Health |
| B | U.S. Should Ratify the U.N.’s Convention on the Rights of the Child - First Focus on Children [Internet]. 2009 [cited 2023 Aug 2]. Available from: <https://firstfocus.org/blog/us-should-ratify-the-uns-convention-on-the-rights-of-the-child> | Grey Literature | Pro | 3 | Child Rights Activist |
| B | Becker J. America Should Not Lag Behind on Protecting Children [Internet]. Human Rights Watch. 2019 [cited 2022 Sep 13]. Available from: <https://www.hrw.org/news/2019/11/18/america-should-not-lag-behind-protecting-children> | Grey Literature | Pro | 9 | Child Rights Activist |
| B | Human Rights Watch [Internet]. 2022 [cited 2022 Sep 13]. How Do US States Measure Up on Child Rights? Available from: <https://www.hrw.org/feature/2022/09/13/how-do-states-measure-up-child-rights> | Grey Literature | Pro | 3 | Child Rights Activist |
| C | Wilkins RG, Becker A, Harris J, Thayer D. Why the United States Should Not Ratify the Convention on the Rights of the Child. St Louis Univ Public Law Rev. 2003;22:411. | Grey Literature | Opposed | 11 | Law |
| C | Kilbourne S. Placing the Convention on the Rights of the Child in an American Context. Hum Rights. 1999;26(2):27–31. | Grey Literature | Pro | 18 | Law |
| C | Caplan AL, Hotez PJ. Science in the fight to uphold the rights of children. PLOS Biol. 2018 Sep 18;16(9):e3000010. | Medline | Pro | 6 | Medicine/ Public Health |
| C | Kitts J, McDonald K. Battle over Children’s rights at the United Nations Special Session on Children. Paediatr Child Health. 2002 Nov;7(9):617–8. | Grey Literature | Pro | 5 | Law |
| C | Mayer A. Reflections on the Proposed United States Reservations to CEDAW: Should the Constitution Be an Obstacle to Human Rights. UC Law Const Q. 1996 Jan 1;23(3):727. | Grey Literature | Pro | 4 | Law |
| C | Melish T. From Paradox to Subsidiarity: The United States and Human Rights Treaty Bodies. Yale J Int Law. 2009 Jan 1;34:389–462. | Grey Literature | Pro | 12 | Law |
| C | Getgen JE, Meier BM. Ratification of human rights treaties: the beginning not the end. The Lancet. 2009 Aug 8;374(9688):447–8. | Grey Literature | Pro | 3 | Medicine/ Public Health |
| C | HuffPost [Internet]. 2014 [cited 2022 Aug 4]. Why Won’t the US Ratify the UN’s Children’s Rights Convention? Available from: <https://www.huffpost.com/entry/why-wont-the-us-ratify-th_b_6195594> | Grey Literature | Pro | 15 | Child Rights Activist |
| C | Abaya M, Todres J. US High-Level Office for Children is Critical for Children’s Rights [Internet]. Health and Human Rights Journal. 2022 [cited 2022 Aug 4]. Available from: <https://www.hhrjournal.org/2022/03/us-high-level-office-for-children-is-critical-for-childrens-rights/> | Grey Literature | Pro | 3 | Child Rights Activist |
| C | The United States has not ratified the UN Convention on the Rights of the Child [Internet]. [cited 2022 Aug 20]. Available from: <https://atlascorps.org/the-united-states-has-not-ratified-the-un-convention-on-the-rights-of-the-child/> | Grey Literature | Pro | 10 | Child Rights Activist |
| C | American Civil Liberties Union [Internet]. [cited 2022 Aug 13]. There’s Only One Country That Hasn’t Ratified the Convention on Children’s Rights: US. Available from: <https://www.aclu.org/blog/human-rights/treaty-ratification/theres-only-one-country-hasnt-ratified-convention-childrens> | Grey Literature | Pro | 9 | Child Rights Activist |

**Supplement 4: Informant Selection Process**

Interviewed: 13

(6 original invitations and 7 snowball referrals)

Interviewed: 1

No answer or declined: 6

Answered the question by email only (not included): 3

Snowball referral or recommendation: 1

Invitations to child rights advocates, global child health experts, child rights lawyers, child health advocates, Gen Z groups, & politicians via email, Twitter, LinkedIn: 24

Invitations to opposition groups via email or website: 9

No answer or declined: 18

Snowball referral or recommendation: 12

No answer or declined: 5

**Supplement 5: Consent form**

**Title of Project: Global Child Health as a Human Right: Why had the United States not ratified the UN Convention on the Rights of the Child?**

**Name of PI/Researcher responsible for project:	Lia Harris**

| **Statement** | **Please check each box** |
| --- | --- |
| I confirm that I have read the information sheet dated June 2023 (version 1.2) for the above-named study. I have had the opportunity to consider the information, ask questions and have these answered satisfactorily.  **OR**  I have had the information explained to by study personnel in a language that I understand. I have had the opportunity to consider the information, ask questions and have these answered satisfactorily. | ☐ |
| I understand that my participation is voluntary and that I am free to withdraw at any time without giving any reason, without my medical care or legal rights being affected. | ☐ |
| I understand that information I provide during the study may be looked at by authorised individuals from London School of Hygiene and Tropical Medicine, where it is relevant to my taking part in this research. I give permission for these individuals to have access to my records. | ☐ |
| I understand that data about/from me/the participant may be shared via a public data repository or by sharing directly with other researchers, and that I **will** be identifiable from this information.  **OR**  I will verbally request that my data remain anonymous at any time before, during, or after my participation. | ☐ |
| I agree to take part in the above-named study | ☐ |

Printed name of participant	Signature of participant	Date

(please type your name)

Printed name of person obtaining consent	Signature of person obtaining consent	Date
